# Traditional Healers as Client Advocates in the HIV-endemic Region of Maputo, Mozambique: Results from a qualitative study

**DOI:** 10.1101/19008490

**Authors:** Radhika Sundararajan, Patrício V. Langa, Trisha Morshed, Sandra Manuel

**Affiliations:** Department of Emergency Medicine, Weill Cornell Medicine, New York, USA; Weill Cornell Center for Global Health, New York, USA; Faculty of Education, Universidade Eduardo Mondlane, Maputo City, Mozambique; Department of Emergency Medicine, University of California San Diego, San Diego, USA

**Keywords:** Traditional Healers, Mozambique, social capital, qualitative

## Abstract

Traditional healers are commonly utilized throughout sub-Saharan Africa instead of – and in concert with – biomedical facilities. Traditional healers are trusted providers and prominent community members, and could be important partners in improving engagement with HIV services in endemic contexts. Our study sought to understand the roles of healers in the urban setting of Maputo, Mozambique, where HIV prevalence is high and testing rates are low. Qualitative data were gathered through minimally-structured interviews with 36 healers. Analysis followed an inductive, grounded theory approach. Data reveal three themes relevant to improving engagement with HIV services in this endemic region: 1) healers have positive attitudes towards biomedicine; 2) healers advocate for their sick clients; and 3) clients are reticent to present to biomedical facilities. Healers describe their roles as ‘cooperative’ with biomedical providers to provide healthcare for their clients. Results suggest that healers could be considered *critical enablers* to effective HIV programs in communities. They have social and symbolic capital that positions them to beneficially influence clients, and are natural partners for interventions to improve uptake of HIV services.

## Introduction

Throughout sub-Saharan Africa, traditional healers play a central role in communities; the WHO estimates that 80% of the general population of sub-Saharan Africa utilizes these practitioners (World Health Organization, 2002). In Mozambique specifically, they play an even more crucial role in managing health complaints. With independence in 1975 came a massive exodus of Mozambique’s physicians, many of whom were under contract with the Portuguese colonial government. Much of the country’s health infrastructure was additionally decimated by a subsequent 16 year-long civil war. Now, Mozambique faces a dearth of physicians, with only 5 for every 100,000 people (World Health Organization, 2006), among the lowest densities of physicians in the world. Healthcare facilities and healthcare workers are also sparsely distributed around the country, with less than 50% of the population having regular access to health services (Global Health Workers Alliance, 2013); people must travel long distances to access health care at biomedical facilities. There are no official statistics describing the number of traditional healers in the country, but they are unofficially estimated at >80,000 (Green, 1999). The study site of Maputo is the nation’s capital, a city of ∼1.7 million people, home to the nation’s largest hospital (Maputo Central Hospital), and region of Mozambique with the greatest coverage of healthcare facilities (Anjos Luis & Cabral, 2016). However, Maputo is home to a large number of healers, demonstrating that their importance is not limited to contexts with low density of biomedical facilities.

Ethnographic and community-based research from sub-Saharan Africa suggests that healers may be favored over biomedical practitioners because they are considered custodians of indigenous knowledge, and commonly hold positions of authority in their communities (Homsy, King, Balaba, & Kabatesi, 2004). As such, their evaluations and recommendations could have cultural appropriateness that biomedicine may not, considering factors outside of biomedical physiology that may contribute to symptoms, such as ancestral unrest, family conflicts, or social transgressions (Green, 1999; Green, Zokwe, & Dupree, 1995). Prior research has also demonstrated that healers are often preferred due to lack of trust in biomedical providers (Homsy et al., 2004; King & Homsy, 1997), and prior negative experiences at healthcare facilities (Morris, 2001; Sundararajan, Kalkonde, Gokhale, Greenough, & Bang, 2013), including “stock outs” of medications, malfunctioning machines, long wait times to access service, and perceived breaches of confidentiality by biomedical providers.

Utilization of traditional healers is particularly relevant to the Human immunodeficiency virus [HIV] epidemic Mozambique. In 2014, Mozambique was noted to have the 3^rd^ highest rate of AIDS-related deaths in sub-Saharan Africa, behind Nigeria and South Africa (UNAIDS, 2014a). The most recent national survey estimates HIV prevalence among sexually active adults at 10.5%, with 1.5 million people living with HIV/AIDS. In Maputo, prevalence is much higher (16.8%), and is where 9% of all Mozambicans living with HIV reside (PEPFAR, n.d.). Despite this, lifetime HIV testing rates in Mozambique are very low (37% for women, and 19% for men)(Health, 2010). The country’s long civil war has also impacted the current population-level skew that is particularly relevant to the HIV epidemic: 51% of the current population is of sexually active age (Global Health Workers Alliance, 2013). Studies throughout sub-Saharan African have demonstrated that traditional medicine is frequently utilized by HIV-infected patients, both prior to and following diagnosis (Hughes et al., 2012; Moshabela, Pronyk, Williams, Schneider, & Lurie, 2011; Wanyama et al., 2017). Prior research in Mozambique has demonstrated that healer utilization is associated with delays in HIV diagnosis (Audet et al., 2014), though there are no data for urban centers where biomedicine is readily accessible.

Mozambican healers work as spiritualists (*Nhamussoro*), herbalists (*Nhangarume*), or birth attendants. Spiritual healers claim the ability to identify and remove curses, or address ancestral unrest through rituals or incantations. In addition to this specialization, they also utilize herbs to heal (via ingestion, “bath”, or “vaccination” where herbs are rubbed into a small incision made in the skin). While spiritual healers use herbs in their treatments, herbalists deal exclusively with herbal remedies. Birth attendants provide prenatal and obstetrical care to women, and employ ‘traditional’ methods for birthing outside of biomedical facilities.

Traditional healer services are formally endorsed by the Ministry of Health, and are largely overseen by the Association of Traditional Healers [Associação dos Médicos Tradicionais de Moçambique, AMETRAMO]. AMETRAMO-affiliated practitioners have standardized treatments and fees for service, and attend periodic training sessions, in cooperation with the Ministry.

Little is known about healers’ practices in urban settings, engagement with biomedical structures, and HIV-relevant practices. This qualitative study addresses this gap in knowledge by 1) considering content and sources of HIV-related knowledge among traditional healers in Maputo City; 2) characterizing healer attitudes towards biomedicine; and 3) describing actions taken by healers when caring for a client they suspect to be HIV-infected.

## Materials and Methods

### Study Design

We enrolled 36 traditional healers practicing throughout Maputo City District between April and November 2016 [Figure 1]. A qualitative study design was employed, with semi-structured interviews to elucidate current practices and beliefs about HIV, and interfaces with biomedicine. An interview guide was created in English, translated into Portuguese and the local language (*Changana*), and back-translated into English to verify preservation of meanings. All participants provided socio-demographic information, including practice specialty, years of practice, highest level of education, client volume, and income. The interview guide was piloted with two traditional healers in March 2016, whose responses are not included in this analysis.

**Figure.**
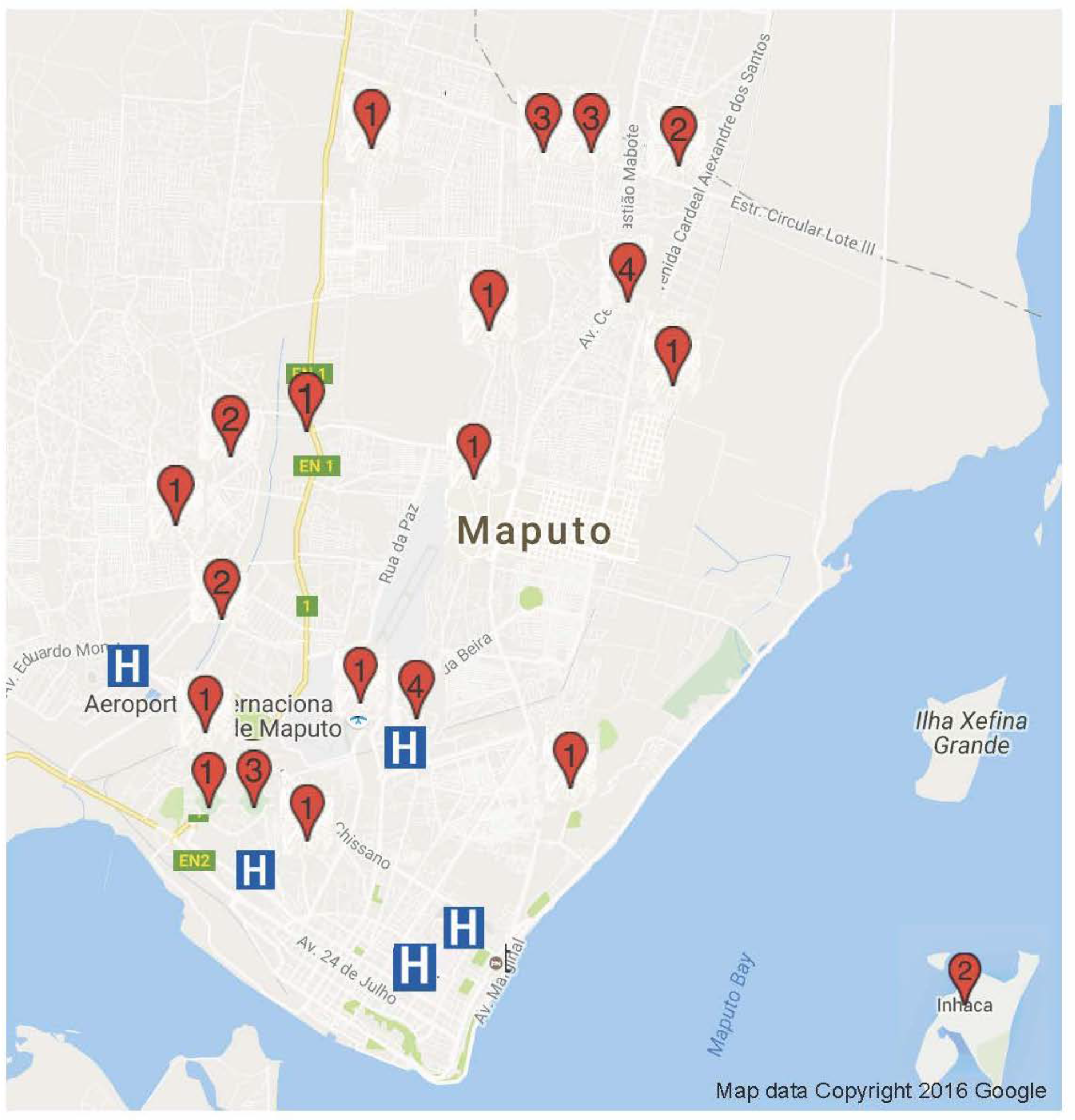
Map of participant practice locations around Maputo City. Red pin locations and numbers indicate location and number of participants from this neighborhood. Blue “H” demonstrates location of hospital. Map Data: © 2016 Google. The street map and hospital locations are freely available on maps.google.com. Permission was not required to reproduce this map image.

### Participant Recruitment and Sampling

During recruitment, research assistants introduced themselves and the study as affiliated with the local university; care was taken not to represent the study or its staff as representatives of hospitals or clinics. Approximately half of our participants (N=15) were recruited in cooperation with AMETRAMO. The remaining participants (N=21) were recruited with assistance from healers and community members, to avoid including only healers who were supportive of the AMETRAMO organization.

We employed a purposive sampling strategy to identify “information rich” cases to maximize the quality of our qualitative data: potential participants having knowledge and experience with our topics of interest (Palinkas et al., 2015). Sample size of N=36 was guided by the concept of qualitative *data saturation* where interviews no longer reveal substantially new or relevant content, and categories are sufficiently developed based on completed interviews (Corbin & Strauss, 2014). Transcripts were analyzed throughout the data collection period; iterative engagement with the dataset allowed timely communication among the research team to direct interviews ensuring emerging themes could be explored and well developed.

### Data Collection

Healers were eligible for participation if they were 1) 18 year of age or older; recognized by the community as a practitioner of traditional medicine; and 3) able to provide informed consent in Portuguese or Changana. Semi-structured interviews lasted approximately 1 hour, and were conducted by four Mozambican research assistants (two female, two male) with educational backgrounds in behavioral science, and fluent in English, Portuguese and Changana. Research assistants used an interview guide to ensure that the following core topics were discussed consistently: 1) HIV knowledge (symptoms, modes of transmission, risk factors, and diagnosis/treatment/cure) and sources of knowledge; 2) prior experiences and attitudes towards biomedicine; and 3) current traditional treatments provided to clients. Research assistants were also encouraged to explore novel or interesting topics that arose in the course of the interview.

### Data Management and Analysis

Interviews were transcribed by the interviewer into Portuguese (translated if the interview was conducted in *Changana*), and then translated into English. One author (PVL) ensured validity of Changana, Portuguese and English translations through random spot-checking, as well as targeted review of transcripts (when English translation was unclear or confusing, for example), against interview audio. Two authors (RS and TM) reviewed all English transcripts for content relevant to HIV-related knowledge and practices among traditional healers, within 1 week of interview completion. The first author analyzed interview data using an iterative, inductive, grounded theory approach (Corbin & Strauss, 2014). Through open coding, preliminary categories were formed. These categories were reviewed, sharpened, refined through data triangulation, and grouped, with generation of a final set of codes. At the completion of data collection, all transcripts were re-reviewed applying the final set of codes to identify major themes illustrating HIV-related knowledge and practices among healers in Maputo, Mozambique. Example(s) from our dataset are presented in the results section that best illustrate these themes. NVIVO 10.0 (QSR International) was used for qualitative data management, but not in the creation of codes. Demographic characteristics of participants are summarized with basic descriptive statistics.

### Ethical Considerations

This research was approved by the Institutional Review Board of the University of California, San Diego (Protocol #150062) and the Biomedical Ethics Committee of the Universidade Eduardo Mondlane Faculty of Medicine. All participants were volunteers, and provided with informed consent, both written and reviewed verbally to ensure comprehension among those participants with limited literacy. Healers were remunerated with 1000 Meticais (∼$15 USD) as compensation for time lost to revenue generation. This remuneration rate was set by AMETRAMO, and is the standard rate for members participating in research studies. The same remuneration was extended to non-AMETRAMO participants in the interest of equity. All audio and data files were stored in password protected electronic files, accessible only to study staff.

## Results

### Characteristics of participants

Table 1 shows characteristics of our participants. In our sample, the vast majority self-identified as spiritual healers (N=34, 94%) and two identified as purely herbalists. There were no birth attendants in our sample, potentially due to proximity of biomedical facilities for prenatal/obstetrical services. All 36 healers approached for enrollment agreed to participate in the study. Consistent with other research involving healers in Mozambique and South Africa, our participants are largely female (67%).

**Table 1.**
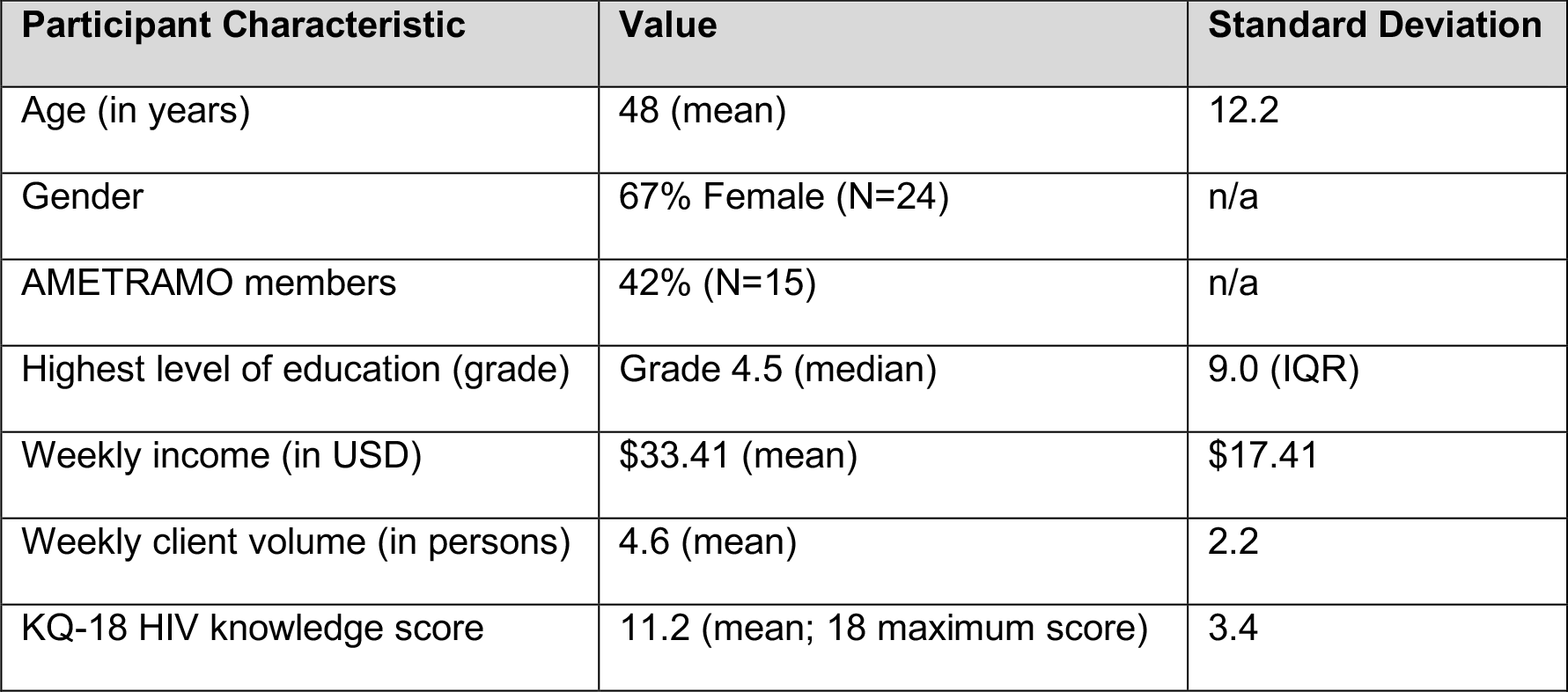
Characteristics of study participants (N=36)

### Interview Themes

We note three major themes within our interviews that are relevant to understanding the roles of traditional healers in this highly HIV-endemic context: 1) praise for biomedicine; 2) advocacy for sick clients; and 3) client reticence for biomedical testing.

### Praise for Biomedicine

Among our sample, healers report uniformly high praise for biomedicine and biomedical providers. Participants describe biomedicine as “curing”, and “as important” as traditional medicine in its healing properties. All participants report use of hospitals and clinics when they and their families fall ill. Specifically, participants admire biomedicine for its ability to “look within” for “internal” maladies through use of serum testing and radiographs, while traditional approaches cannot. Some describe biomedicine as “complementary” to traditional healing, and themselves “collaborators” with biomedical providers. One participant describes a common process of clients tacking between the two realms:

> For the natural ones [biomedical illnesses], I tell the patient to go to hospital. But it is important to say that there are some cases in which the patient had been to the hospital and the disease was not diagnosed. So when they come to me, I remove the evil spirit that was preventing the diagnosis of the disease, and then I send back the patient to the hospital. I also encourage the patient by saying that s/he will get well soon and what s/he needs to do is to follow the hospital doctor’s instructions (Participant 305, Male, 53 years old).

The healer in this case describes a context of *medical pluralism*, where both traditional and biomedical forms of healing are utilized by the client. He is encouraging the client to adhere to instructions provided by biomedical providers, suggesting that healers do not conceive of hospital or clinics as competitive entities, nor are they distrustful of their recommendations. Rather, they offer support and encourage the client’s biomedical utilization.

### Advocating for their clients

Participants describe behaviors that facilitate client utilization of biomedicine, particularly for those presenting with symptoms of Acquired immunodeficiency syndrome [AIDS]. They report familiarity of symptoms of HIV infection as those related to AIDS (weight loss, chronic cough, alopecia, diarrhea, chronic skin rashes/wounds); these clients are immediately referred to the hospital. Our results support the concept that healers are *advocates* for their clients, and consider themselves to be working in cooperation with biomedical providers. Advocacy behaviors includes escorting patients to the hospital themselves, phoning in “referrals” to hospitals, “begging” or otherwise persuading clients to visit the hospital. Such advocacy is underlain by feelings of “love” and “obligation”, and acting in the best interests of clients, whom they consider “family members”. A healer describes her relationship with one of her clients, who was recently diagnosed as HIV-infected:

> It still secret [that my client is HIV+]. No one else knows apart from me, even his relatives. So now I have become his relative, because I am the one who escorts him to the clinics, a thing which his relatives do not do … When we were at the hospital, the only thing that the patient was saying is that he ‘preferred to die’. Then I asked, ‘how about the two daughters that you have?’ I told him that he is still young and he can live a long life if only he takes care of himself and starts to medicate. It took a lot of effort in order to enroll [in HIV care] at the hospital for him to start medications, and sure enough, I managed to help him do it. He is medicating and now he shows some good signs (Participant 301, Female, 49 years old).

This participant elaborates on a sense of responsibility in convincing a sick-appearing client to go to the hospital:

> He did not welcome the news [of going to the hospital], but since we fight for people’s lives, we need to be bold enough to do so. We cannot mess up with these diseases, especially AIDS … Diseases like this one, they need medical attention (Participant 208, Female, 45 years old).

Another healer describes his role in the lives of his clients: “The job we have is so important, I say. Because from the traditional perspective of healing there is a lot that we are able to help a lot of people with … We are part of what helps a human being” (Participant 202, Male, 59 years old).

### Clients “run” from biomedicine

Traditional healers in our sample report that their clients frequently object to their referrals for biomedical evaluation and testing, and will “run away” from the healer to avoid this outcome. Healers “know by looking at someone” if the client is “sick or not”; sick clients warrant urgent referral to hospitals. However, they have learned to approach these clients “carefully” to avoid causing them to flee:

> We have learned now. We know how to talk to them, because a person in those conditions [when clients are sick] is so sensitive. If we do not attend to him here, he will not even get to a hospital … I will handle [that client] carefully, because you cannot communicate aggressively to the patient. If you do so, he will run away … so, you be tender as you inform him, until he goes to hospital (Participant 203, Female, 50 years old).

Our participants attribute client resistance to fear of biomedical testing (for HIV, specifically). One participant reports, “Many times, people say that when a traditional healer fails to help you, it means you are dead already” (Participant 202, Male, 59 years old). This saying underlies the concept that traditional practitioners are trusted and preferred for their capacity to *heal*, while visits to biomedical facilities may be associated with bad outcomes and death. Another healer suggests that their clients prefer traditional medicine as a means of avoiding their medical diagnoses: “When they come here, sometimes they are avoiding going to hospitals, or they have been there and were diagnosed with HIV and he hides it and come to us, and you ask him back to hospital – again he runs away” (Participant 204, Female, 66 years old). Healers also suggest that clients avoid biomedical facilities due to perceived poor quality of services, such as “long queues” or “defective machines” and the fact that practitioners are “not friendly to patients in our hospital. I think when one thinks of having his HIV status know by those professionals, I thinks it scares him” (Participant 213, Female, 35 years old).

## Discussion

This qualitative study elucidates experiences of healers practicing in Maputo, Mozambique, an HIV-endemic region which is also home to the highest density of biomedical resources in the country (Anjos Luis & Cabral, 2016). Our results demonstrate that Maputo is a medically pluralistic context with cyclical/concurrent traditional and biomedical use within one’s therapeutic itinerary. Participants state that clients “run” from biomedical facilities, potentially due to mistrust of services and providers, and preference for culturally competent traditional healing. Despite low opinions and avoidance by clients, our healer participants report high regard for biomedicine. They consider themselves as collaborators with biomedical providers, and advocate for clients to escalate care, often facing resistance from clients whom they refer to the hospital. This provides a unique, holistic arrangement where biomedical and traditional approaches to healing are accepted and encouraged.

Our research demonstrates that traditional healers function in their communities as client advocates. Advocacy efforts are based on their sense of obligation to clients, whom they consider to be close, like family members. Healers develop strategies to overcome client resistance to HIV testing, by approaching them in a “tender” manner, through developing rapport, and providing reassurance and support. Further, our results describe an important factor contributing to low rates of HIV testing in Mozambique: client reticence for biomedical testing. This describes a bottleneck in the cascade of HIV care. If HIV-infected patients are not identified through voluntary testing, they cannot be enrolled into care. This is particularly problematic because early diagnosis and initiation of ART therapy is crucial to preventing transmission and optimizing health outcomes (The INSIGHT START Study Group, 2015). Our results also speak to the phenomenon of the widening gender gap in life expectancy among HIV-infected populations in sub-Saharan Africa, where men have two-fold higher mortality rates than women (Bor et al., 2015). These ‘missing men’ (Tsai & Siedner, 2015) may be clients who “run away” from hospitals and biomedical facilities. Efforts to improve engagement with male populations in HIV endemic regions should therefore consider those who visit healers, and how collaborative efforts may overcome reticence toward engagement with HIV services. This holistic approach to client health explains why healers maintain positions of authority in their communities. This is no less important in the urban, capital city than it is for clients living a rural district with no local hospital.

Based on our results, we suggest that traditional healers could be considered one of the “critical enablers” (UNAIDS, 2012) of effective responses to the AIDS epidemic in Mozambique, if not throughout much of sub-Saharan Africa where they practice. Critical enablers are stakeholders who can “overcome major barriers to [HIV] service uptake, including social exclusion, marginalization, criminalization, stigma and inequity. Critical enablers are crucial to the success of HIV programs in all epidemic contexts” (UNAIDS, 2012). Our results, and others, indicate that healers are preferred providers due positions of respect and trust in their communities (Green, 1992), their ability to provide holistic and culturally-appropriate care (O’Brien & Broom, 2014; Taylor, Dolezal, Tross, & Holmes, 2008), and are relied upon in regions that lack available biomedical resources (Morris, 2001).

Traditional healers possess *social capital*, amassed through their engagement in “networks of social acquaintance and recognition” (Bourdieu, 1986). These networks of interactions and prestige also generate *symbolic capital*, where “power is granted to those who have obtained sufficient recognition to be in a position to impose recognition” (Bourdieu, 1989). Our data demonstrate how healers mobilize their social and symbolic capital to benefit clients through their advocacy – facilitating access to healthcare resources, and encouraging clients to undergo HIV testing – through a cooperative approach to healing. As such, healers are gatekeepers to communities with poor biomedical engagement. National and international responses to ending the HIV epidemic by 2030 (UNAIDS, 2014b) must consider the impact of these practitioners within their communities. Our data, like others from varying contexts (Audet, Hamilton, Hughart, & Salato, 2015; Homsy et al., 2004; King & Homsy, 1997), demonstrate cooperative approaches to caring for clients, suggesting that healers are willing to work in collaboration with biomedical providers, researchers, and public health officials.

### Limitations

We recognize some limitations of our study. First, qualitative data are not intended to be largely generalizable; rather, they provide detailed and highly contextual information about a population of interest. As such, we acknowledge that our qualitative findings may be specific to a population of traditional healers practicing in a relatively well-serviced city (Maputo, Mozambique). We do, however, note some similarities with studies with healers in more remote area of sub-Saharan Africa, particularly insofar as we describe positive attitudes towards biomedicine and willingness to collaborate with biomedical providers. Second, we did not amass evidence to support the notion that biomedical providers reciprocate the cooperative spirit described by study participants.

## Conclusions

Our data include findings which can inform efforts to meet the Joint United Nations Programme on HIV/AIDS 90-90-90 benchmarks to end the epidemic (UNAIDS, 2014b). We present three important themes relevant to the roles of traditional healers in HIV-endemic regions: 1) positive attitudes towards biomedicine that underlies cooperative healing for their clients; 2) advocacy for clients; and 3) client reticence for biomedical testing. Healers practicing in these regions could be considered *critical enablers* to effective HIV programs. Their social and symbolic capital positions them to beneficially influence clients. They are natural partners to improve uptake of HIV services in communities with poor biomedical engagement.

## Data Availability

Data requests will be considered on a case-by-case basis by contacting the first author directly.

## Acknowledgements

The authors are grateful to Ms. Olga Tomas, AMETRAMO President, for her support and assistance with member participant recruitment. We are also grateful to our Research Assistants for their hard work carrying out the data collection: Felício Almeida, Alberto Manhuja, Andreia da Silva, Delfinada Silva. We also wish to extend our sincere gratitude to the Traditional Healers who participated in our research, and for sharing their experiences and insights with us.

## Authors’ contributions

RS conceived and designed the study, primarily conducted data analysis, and drafted the manuscript. PVL and SM contributed to study design, oversaw data acquisition, contributed towards data analysis, revised the manuscript for critically important intellectual content. TM oversaw data acquisition and revised the manuscript for critically important intellectual content. All authors approve of the version of the manuscript for submission.

